# Use of cranial ultrasonography to improve prompt and early diagnosis of meningitis at the Neonatal Centre of Excellence, University Teaching Hospitals, Lusaka, Zambia

**DOI:** 10.64898/2026.07.29.26359192

**Authors:** Ogah Adenike, David Hamer

## Abstract

**Background:** Despite cerebrospinal fluid (CSF) analysis being the gold standard for definitively diagnosing meningitis, its practical application has presented considerable difficulties, especially in environments with limited resources. Cranial ultrasound (CUS), while not a replacement for CSF analysis, provides rapid imaging to identify meningeal irregularities. Nevertheless, achieving a timely diagnosis of meningitis, particularly in its nascent stages, remains problematic, and healthcare professionals exhibit a notably low awareness of CUS’s utility in diagnosing this condition.

**Methods:** This was a prospective cohort that recruited 273 term mother-neonate pairs. The neonates were initially divided into those with sepsis-meningitis (exposure group) and those with sepsis (non-exposure group) based on clinical assessment done by the admitting team. The research team further divided the participants into 4 diagnostic subgroups based on CUS findings: positive CUS for meningitis, positive clinical diagnosis for meningitis, positive CUS & clinical diagnosis for meningitis and those with negative CUS & clinical diagnosis for meningitis /sepsis only. Data on socio-demographics, clinical characteristics, blood works and CSF analysis reports, neurologic deficits and mortality outcomes were recorded for each neonate. Descriptive and inferential statistics were performed.

**Results:** The 4 diagnostic subgroups based on CUS findings were: positive CUS for meningitis (24.5%), positive clinical diagnosis for meningitis (4.4%), positive CUS & clinical diagnosis for meningitis (6.6%) and those with negative CUS & clinical diagnosis for meningitis /sepsis only (64.5%). Overall, duration of hospitalization was 11 days (range 2-45 days; interquartile range [IQR] 6,16) and the median chronological age of the neonates was 13 days (IQR 7, 21). Meningitis was suspected in 11% of neonates admitted with clinical sepsis. Uptake for lumbar puncture (LP) or ventricular tap (VT) was low at 4.4% (n = 12), underlining barriers to CSF-based diagnosis in this setting. Late-onset sepsis was associated with only 31.9% of NSM. The chronological age of the neonates at admission (p=0.002) and their duration of hospitalization (p=0.042) were significantly different across the 4 categories of neonates. Prominent sulci, hyperechoic brain lesions, ventriculitis, and lateral ventriculomegaly were the most common abnormal findings on CUS. Overall, neonates with meningitis presented later in age and stayed longer on the ward than those with sepsis: clinical meningitis diagnosis was likely to be made in older neonates and was associated with a shorter duration of hospitalization than CUS-diagnosed meningitis. Overall mortality rate was 2.2%. Mortality (16.7%) was highest amongst those with clinical & CUS diagnosed-NSM. Prevalence of Near-Miss cases of NSM was at least 27.6%. A neonate with clinical diagnosis of NSM was 3.94 times (95% CI 1.80, 8.62; p<0.001) more likely to have abnormal CUS at the time of admission. Percent agreement between the clinical and CUS diagnosis for NSM was 71.1%. Nine (75%) out of the 12 CSF reports was positive for meningitis; and the majority 55.6% (5) of the positive CSF report belonged to the category of neonates with sepsis only. Compared to CSF analysis, sensitivity of clinical diagnosis of NSM was 22.2%; specificity was 33.3%; positive predictive value (PPV) was 50%; negative predictive value (NPV) was 12.5% and likelihood ratio (LR) was 0.33. Whereas, sensitivity of CUS was 44.4%; specificity of CUS was 66.7%; PPV was 80%; NPV was 28.6% and LR was 1.33.

**Conclusion:** The CUS was only moderately effective at diagnosing meningitis whereas the presence of positive CSF among neonates with sepsis only within this study, further reaffirms the irreplaceability of CSF analysis in the diagnosis of NSM. Nevertheless, the integration of clinical assessment and CUS findings for diagnosing NSM emerged as possessing greater clinical significance in contexts characterized by limited resources. The systematic adoption of CUS for neonates exhibiting features suggestive of sepsis or meningitis (while exploring measures to improve on uptake of CSF analysis) could enhance the promptness of diagnosis, inform the selection of suitable therapeutic interventions, and potentially mitigate mortality rates and the occurrence of long-term neurological impairments, especially within environments facing resource limitations.

## Introduction

Neonatal sepsis-meningitis (NSM) is a diffuse inflammation of the meninges during the first 28 days of life, that is usually due to acute bacterial infection.^1^ Meningitis is almost always a result of hematogenous spread of pathogens into the central nervous system (CNS) through the choroid plexus and then disseminates via the cerebrospinal fluid (CSF) in the ventricles. Choroid plexitis, ventriculitis, arachnoiditis, vasculitis, cerebral edema and encephalopathy are detectable on CUS.

To exclude the diagnosis of meningitis, all these parameters: CSF culture, gram stain, chemistry (glucose and protein) and cytology or cell count must be normal.^2,3^ Hence, the gold standard for confirming meningitis is CSF analysis. Despite advances in the management of NSM, its case fatality rate remains high between 13-59%,^2^ and there is a marked and permanent neurologic sequelae seen in 20-58% of the cases, which is unacceptably high, especially in developing countries.^4,5^ Currently, there is no country-specific data from Zambia about the burden of NSM. Most estimates are inferred from nearby African countries.^6^ In an institutional-based cross-sectional study (2021) in Ethiopia, a neighboring country to Zambia, the prevalence of NSM was 19.3% among neonates with suspected sepsis; 22.8% in early-onset neonatal sepsis (EONS), and 16.8% in late-onset sepsis (LONS). Although, NSM is documented in literature, to be more common in LONS than EONS. Suffice it to say that, the neonate remains the most vulnerable to meningitis in all stages of human life.^7^

In the context of this publication, early-onset sepsis is characterized by the manifestation of septic symptoms within the initial seven days postnatally, following a period of discharge from the health facility. Conversely, late-onset sepsis denotes the onset of sepsis occurring subsequent to the first week of life.^8^

In a setting, where access to CSF evaluation, for confirming meningitis or other laboratory support is limited, the chances are very high that NSM could be missed; where the presentation is often subclinical and non-specific and may not even be accompanied with sepsis. Despite being irreplaceable, lumbar puncture (LP), an invasive procedure, to obtain CSF, may be contraindicated in very sick neonates and CSF analysis-unrevealing in cases of ventriculitis and brain abscess, as these require direct imaging, which is the case with CUS.^7^ CSF culture may also be negative, if the neonate has been exposed to intrapartum or empirical antibiotics before presenting at the neonatal unit or had viral NSM.^9, 10^ Molecular testing of CSF is only available in very few resource-limited settings. In certain environments, antigen testing is conducted using a restricted selection of organisms, whereas in high-income nations, multiplex polymerase chain reaction (PCR) is employed for the detection of numerous pathogens responsible for meningoencephalitis.^8^ Infants can develop hydrocephalus even after standard meningitis treatment and with normal cerebrospinal fluid (CSF) because ventriculitis and choroid plexitis, sources of recurring infections,^11^ are hard to treat. Earlier detection of ventriculitis with CUS would require extended antibiotic treatment and follow-up beyond typical NSM care. While ventriculitis and ventriculomegaly can be reversed if caught early, this chance is often missed in practice. In a 2019 study in Zambia, post-infectious hydrocephalus accounted for 65% of 378 pediatric hydrocephalus cases at Beitcure and University Teaching Hospitals.^6^ This increase in hydrocephalus and cerebral palsy cases at UTH, averaging 24 per week, may stem from untreated or poorly managed NSM.

To mitigate these challenges, an alternative to LP/VT, to strengthen the suspicion of CNS infection, must be sought. Literature has confirmed the critical role of CUS, a non-invasive, low-cost, portable, easy, speedy and relatively harmless procedure, with no need for sedation, and its lack of ionizing radiation makes it superior to other cross-sectional imaging, such as CT scan and MRI. Hence, CUS can facilitate early diagnosis, prompt appropriate treatment, guide in the duration of treatment, early recognition of complications of neonatal meningitis^10^ and potentially reduce adverse long-term neurological outcomes.

CUS can be repeated as often as necessary to produce serial images during the course of the disease. The portability of CUS is of utmost importance in the evaluation of very sick neonates that cannot be moved, as a screening and diagnostic test. CT scan and MRI services are beyond the reach of majority of health facilities in developing countries.^10^

Even though there is no contraindication to this form of imaging, CUS may not detect any abnormality in some early mild cases of NSM, some abnormal findings may be transient, and some may be subjective depending on the level of competence and experience of the sonographer and some findings may be shared by other diagnosis.^10^

Even in affluent countries, only 30 to 50% of cases of NS in the neonatal intensive care unit (NICU) receive LP/VT, underestimating the incidence of NSM.**^12^** In low- and middle-income countries (LMICs), CUS has been employed for monitoring long-term consequences of meningitis, including hydrocephalus; however, its application in diagnosing NSM has been rare.

Locally, the challenges of acquisition of parental/caregiver’s consent for LP/VT procedures, coupled with constrained resources and subsequent delays that precipitate complications, and healthworkers’ unawareness of the role of CUS in NSM diagnosis, underscores the necessity for an unobtrusive methodology.

Therefore, this study aimed to determine the proportion of septic neonates that have CNS infections using CUS; to determine the proportion of near-miss cases of NSM at the time of admission at UTH and the utility of CUS versus clinical assessment of NSM, to document the early and late CUS findings of NSM, and the short-term outcome of NSM i.e. neurologic deficits, discharge or death.

## Methods

### Study Design

This was a diagnostic yield study with an initial 2-arm prospective cohort design -the exposure and non-exposure groups.

### Study site and Participants

Two hundred and seventy-three eligible term mother-neonate pairs were recruited and divided into 2 broad groups. The exposure group included neonates/infants with suspected sepsis-meningitis (NSM), 30 in number; while the non-exposure group were neonates/infants with suspected sepsis (NS), 243 in number; based on clinical evaluation at the time of hospitalization by the admitting team of doctors on duty. The research team had no influence on the initial grouping of participants in the study. CUS was performed by the Principal Investigator at the point of recruitment into the study for all the 273 participants. After the CUS, the participants were further divided into 4 diagnostic subgroups. These 4 subgroups were those with: i) positive CUS-diagnosed NSM, ii) positive clinically-diagnosed NSM, iii) positive CUS- & clinically-diagnosed NSM and those with iv) negative CUS-& clinically-diagnosed NSM (Sepsis only sub-group). Once, meningitis was suspected, whether clinically or by CUS the patient’s treatment plan was updated to conform to NSM protocol and reviewed later with CSF analysis report.

#### Inclusion and Exclusion criteria

All infant admissions, chronologically aged 0-3months with clinical diagnosis of sepsis at the Neonatal Centre of Excellence of UTH-Children Hospital, Lusaka, Zambia. Babies with obvious brain malformation were excluded from the study.

#### Sample Size and Sampling Strategy

Given the available evidence from literature,**^13^** the assumption was that abnormal CUS in NSM was approximately 81.7%.^3^ We expected to observe a 63% difference in abnormal CUS prevalence rate between the exposed and unexposed groups. Considering odds ratio of 20, risk ratio of 4.5, 80% statistical power, two sided confidence level of 95% and using likelihood-ratio test for comparing two independent proportion gives a minimum sample size of 24 (12 per group) to be sufficient to determine the hypothesized difference.**^14, 15^** However, sample size of 273 was utilized for robust regression analysis. Sampling strategy was consecutive, on a first-come-first-serve basis until sample size was completed. Recruitment of patients into the study took place only after the admitting Doctors have made their clinical diagnosis, commenced initial treatment and the research team had obtained informed consent from the caregivers.

### Research Outcomes and Measures

#### The dependent variables

The primary dependent variable was CUS findings at recruitment (normal or abnormal) and secondary dependent variable was the outcome (death or discharged) at the point of exit from the study. The near-miss cases of NSM in this study, were those, whose admission diagnosis of neonatal sepsis was changed to suspected meningitis, only after documenting abnormal CUS findings and before CSF results were obtained. Whenever NSM was diagnosed during hospitalization, anti-meningitis antibiotics treatment was commenced immediately after collecting the blood and CSF samples.

Clinical diagnosis of NSM was made by the admitting team of Doctors independent of the Research team. The CUS-based diagnosis of NSM was made, when any abnormal CUS signs suggesting meningitis was present during the study.^3,10,^ **^15, 16, 17, 18, 19^**

### The independent variables

The biodata of the parents and patient (neonate/infant), and clinical findings in the history taking, physical examination and laboratory reports of the patient at recruitment, during hospitalization and at exit were documented. Patient’s corrected postnatal age, sex; birth and current-weights, lengths and head circumferences and their z-scores were also recorded.^20, 21^

### Data Collection and Management

The CUS was performed using a full-sized MindRay Consona N6 ultrasound machine equipped with neonatal brain software and probes. The CUS findings on the brain surface, brain parenchyma, ventricular system, measurements of the brain structures and Doppler patterns were recorded and interpreted for each infant. Data collection was obtained using Kobocollect app and transported to Excel spreadsheet. Frozen photos of CUS views were stored in external hard disk drives. Data collection was for 8 months from 4^th^ of August 2025 to 31^st^ of March 2026.

### Statistical Analysis Plan

Data were analyzed using SPSS version 26. Frequencies and percentages were used to report prevalences and describe the CUS findings in the study. Parametric tests (when data was normal in distribution) such as T-test and ANOVA and non-parametric tests (when data was significantly skewed in distribution or too small in size) such as Kruskal Wallis Test were used to assess relationship between the variables. The reporting in this study were guided by the STROBE guidelines for observational studies.^21^

### Ethical/Regulatory Approvals and Consideration

Ethical clearance was obtained from the University of Zambia Biomedical Research Ethics Committee and National Health Research Authority with reference number UNZA-5664/2024. Permission to collect data was obtained from the Office of the Senior Medical Superintendent and Ward Manager of the NCOE at the University Teaching Hospital-Children Division. Written informed consents were obtained from the caregivers to participate in the study. The ALARA precaution was observed throughout the study.

## Results

### Demographic and Clinical Characteristics of the Neonates in the Study

Two hundred and eighty-four mother-neonate pairs were recruited. Data that belonged to 11 participant pairs were removed from the data analysis because of incomplete socio-demographic and laboratory entries.

Female neonates were 45.1% out of the 273 neonatal data analysed. Prevalence of suspected clinical NSM was 11% (30/273). Prevalence of CUS-NSM among suspected clinical NSM was 60% (18/30). Overall, mean maternal age was 28.49years (sd 6.72) and paternal age was 33.95years (sd 7.82). There was no significant difference in the parental ages across the 4 categories of neonates-maternal (p=0.331) and paternal (p=0.472).

Overall, mean gestational age and weight at birth were 38.2 weeks (sd 2.68) and 2.9kg (sd 0.63), respectively; at admission-weight 3.1kg (sd 0.95); length 49.7cm (sd 4.1); head circumference 35.3 (sd 1.47); hemoglobin 13.3g/dl (sd 2.95); oxygen saturation 93.8% (7.39); heart rate 150bpm (sd 21.9); Temperature 37°Celsius (1.28); capillary refill time 2.2sec (sd 0.75) and median pain scale 2 (IQR 1,3).

Overall median duration of hospitalization was 11days; ranging from 2-45days; IQR 6,16. Overall median random blood sugar level at admission was 5mmol/l; ranging from 1-97mmol/l; IQR 4, 6mmol/l. Six (2.2%) out of the 273 neonates died, Table 1. Clinical diagnosis and CUS of each neonate were compared.

**Table 1:**
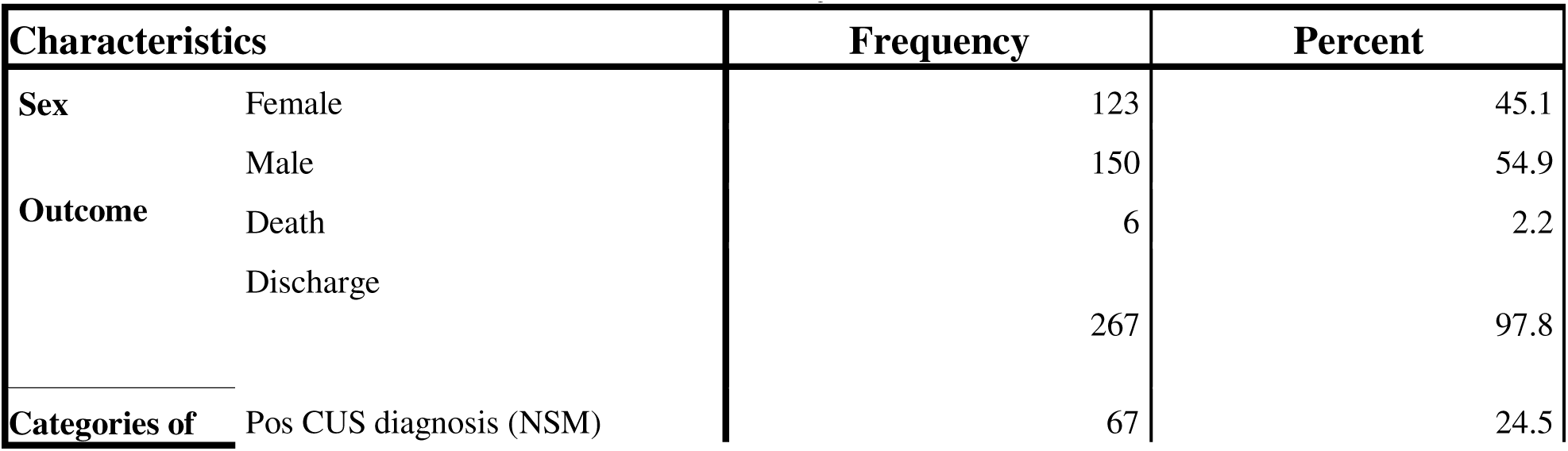
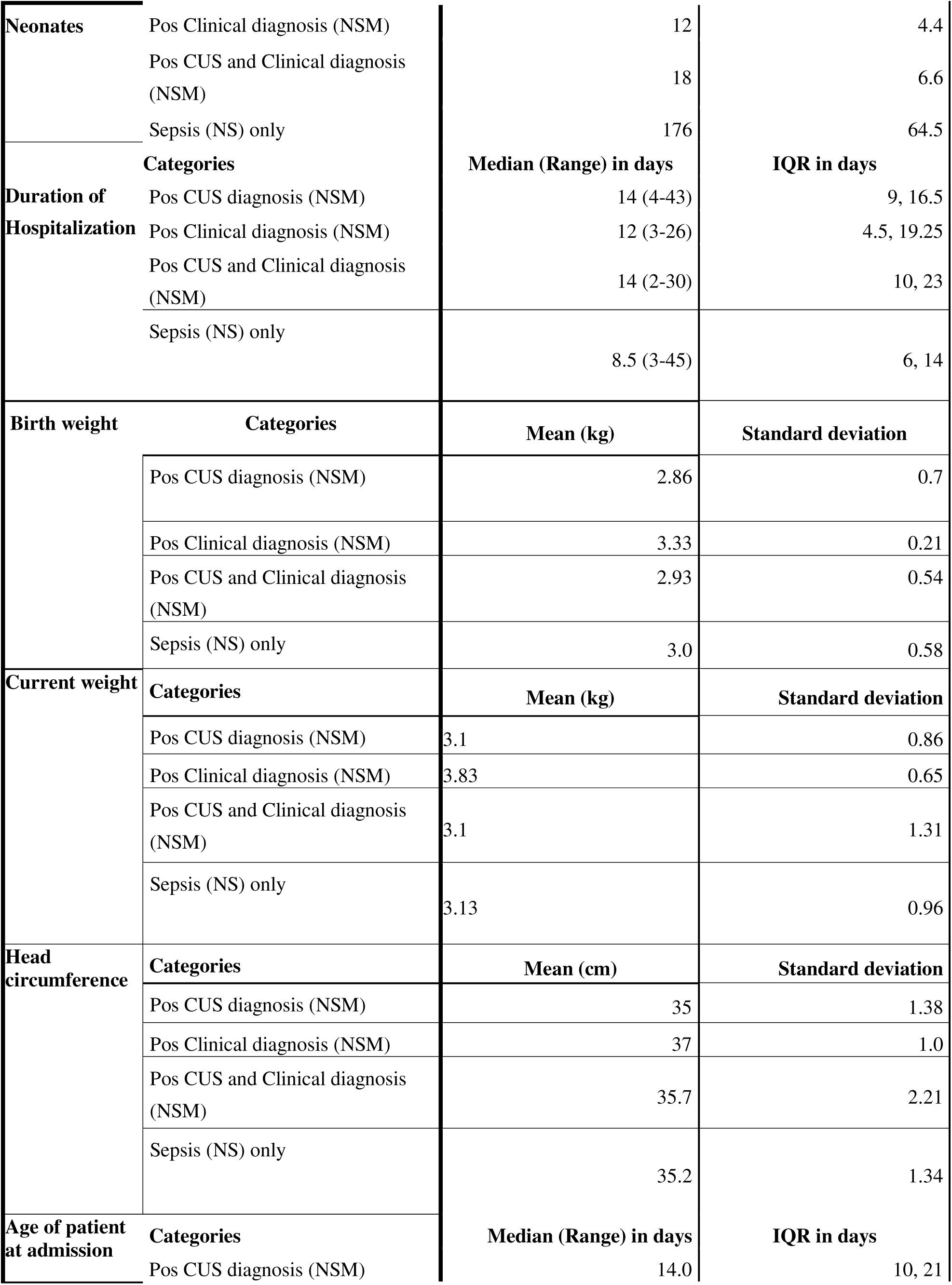
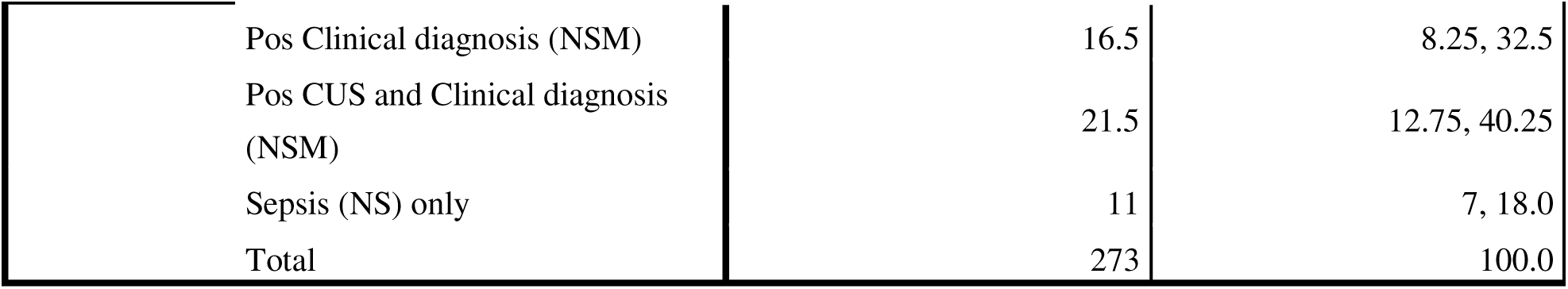
Characteristics of Neonates in the study, n=273.

There was no significant difference in the median random sugar (p=0.201) and pain scale levels (p=0.178) at admission; mean gestational age at birth (p=0.138), mean anthropometric, hemoglobin and vital signs values (p>0.05) across the 4 categories of neonates in the study. However, the age of the neonates at admission (p=0.002) and their duration of hospitalization (p=0.042) were significantly different across the 4 categories of neonates in the study, Table 1.

Though the differences were not statistically significant, the category of neonates with positive CUS & clinical diagnosis of NSM had the lowest hemoglobin and oxygen saturation levels at 11.4g/dl (sd 2.46) and 92.7% (sd 5.9); but the highest heart rate and pain scale at 162.4bpm (sd 18.7) and 4, respectively. The sepsis only subgroup had the highest Hb and oxygen saturations at 13.9g/dl (sd 2.98) and 94.8% (sd 6.26); and lowest pain scale at 2 points, respectively. Of note the subgroup with clinical diagnosis of NSM had the lowest body temperature at admission at 35.7oC (sd 0.58). The head circumference of neonates with positive CSF findings for NSM were significantly smaller by 1.79cm (95% CI -3.37, -0.21) than those with normal CSF, p=0.03.

#### CUS Findings in the study

In Table 2, prominent sulci (17.9%) were the most common abnormal cranial ultrasound finding in the study, followed by hyperechoic brain lesions (13.6%), ventriculitis (11%) and lateral ventriculomegaly (8.4%).

**Table 2:** Cranial ultrasound findings in the study.

| # | Cranial ultrasound findings | Frequency | % |
| --- | --- | --- | --- |
| 1 | Prominent sulci | 49 | 17.9 |
| 2 | Hyperechoic brain parenchyma lesions | 37 | 13.6 |
| 3 | Ventriculitis | 30 | 11 |
| 4 | Lateral ventriculomegaly | 23 | 8.4 |
| 5 | Intraventricular hemorrhage | 21 | 7.7 |
| 6 | Cysts/Cavities in the brain parenchyma | 13 | 4.8 |
| 7 | 3<sup>rd</sup> ventriculomegaly | 11 | 4.0 |
| 8 | 4<sup>th</sup> ventriculomegaly | 10 | 3.7 |
| 9 | CSF debris | 10 | 3.7 |
| 10 | Wide Interhemispheric Fissure | 8 | 2.9 |
| 11 | Extracerebral collections | 6 | 2.2 |
| 12 | Brain parenchyma abscess | 4 | 1.5 |
| 13 | Blurred brain grey-white matter demarcation | 4 | 1.5 |
| 14 | Septae in the CSF space | 3 | 1.1 |
| 15 | White brain | 2 | 0.73 |
| 16 | Slit ventricles | 2 | 0.73 |
| 17 | Wide Cisterna Magnum | 2 | 0.73 |
| 18 | Interhemispheric Fissure shift | 1 | 0.4 |
| 19 | Lenticulostriate vasculopathy | 1 | 0.4 |
| 20 | Brain malformation | 1 | 0.4 |

### Relationship between Clinical and CUS Diagnosis of Neonatal Meningitis in the Study

Thirty (11%) neonates were diagnosed with suspected clinical meningitis at admission, while 85 (31.1%) out of the 273 neonates had abnormal CUS findings suggesting meningitis, Table 3. **Prevalence of Near-Miss cases** of NSM was 27.6% (67), Table 3, p<0.001 (Chi 13.096). Odds of a clinical NSM having abnormal CUS was 3.94 (95% CI 1.80, 8.62).

**Table 3:** Crosstabulation of Clinical Diagnosis and Cranial ultrasound Diagnosis.

|  |  | Cranial Ultrasound Diagnosis |  | Total (%) |
| --- | --- | --- | --- | --- |
|  |  | NSM (%) | NS (%) |  |
| Clinical | Neonatal Meningitis (NSM) | 18 (60) | 12 (40) | 30 (100) |
| Diagnosis | Neonatal Sepsis (NS) | 67 (27.6) | 176 (72.4) | 243 (100) |
| Total |  | 85 (31.1) | 188 (68.9) | 273 (100) |

**Percent Agreement** between the 2 (Clinical and Cranial Ultrasound) diagnosis was calculated as follows: Percentage of the Total (273) concordant for NSM was 6.6% (18/273); while percentage of Total (273) concordant for NS was 64.5% (176/273) between the 2 diagnostic methods. Hence, **Percent Agreement** between Clinical diagnosis and CUS diagnosis of NSM was 64.5+6.6=71.1%. Kappa Statistics as a measure of agreement between Clinical Diagnosis and CUS diagnosis was statistically significant (p<0.001), but at a low value of 0.180, suggesting only slight agreement.

### Cerebrospinal Fluid Findings and Utilities of Clinical Diagnosis and CUS Diagnosis

Only 12 (4.4%) out of the 273 mothers, consented to a lumbar puncture/ventricular tap for their neonates. Two of these CSF samples were straw colored, 3 were cloudy, 2 were blood stained and 5 were clear. Only CSF appearance, chemistry and cytology were documented. CSF culture yielded no organism in all the 12 CSF samples. However, in one of the cloudy CSF samples, *Toxoplasma Gondii* was detected by CSF serology. Ten (83.3%) out of the 12 neonates with documented CSF results were referred to UTH from other health facilities. In Table 4, 9 (75%) out of the 12 CSF reports was positive for meningitis; and a higher percentage 41.7% (5) of the positive CSF report belonged to the category of neonates with sepsis only.

**Table 4:** CSF findings across the categories of Neonates.

|  |  | CSF VERDIT |  | Total (%) |
| --- | --- | --- | --- | --- |
|  |  | Pos (%) | Neg (%) |  |
| Categories of Neonates | Pos CUS and Clinical Diagnosis (NSM) | 2 (66.7) | 1 (33.3) | 3 (100) |
|  | Pos CUS diagnosis (NSM) | 2 (100) | 0 (0) | 2 (100) |
|  | Pos Clinical diagnosis (NSM) | 0 (0) | 1 (100) | 1 (100) |
|  | Neg CUS and clinical diagnosis (NS) | 5 (83.3) | 1 (16.7) | 6 (100) |
| Total |  | 9 (75) | 3 (25) | 12 (100) |

#### Utility of Clinical Diagnosis

Of note, 50% of those with clinical diagnosis of NSM, were confirmed with abnormal CSF findings, while 87.5% of those with clinical diagnosis of sepsis, actually had abnormal CSF findings, Fisher’s Exact Test =0.236. Kappa value was -0.286, p=0.157. OR 0.143 (95% CI 0.008, 2.52). Sensitivity of clinical diagnosis was 2/9=22.2%. Specificity was 1/3=33.3%. Positive Predictive value was 2/4=50%. Negative predictive value was 1/8= 12.5%. Likelihood Ratio was 0.33.

#### Utility of CUS

In contrast, eighty percent of those with abnormal CUS diagnosis were also confirmed to have abnormal CSF findings. Fisher’s Exact Test =1.000. Kappa value was 0.077, p=0.735. OR 1.60 (95% CI 0.10, 24.7). Sensitivity of CUS is 4/9=44.4%. Specificity of CUS is 2/3=66.7%. Positive Predictive value is 4/5=80%. Negative predictive value is 2/7=28.6%. Likelihood Ratio is 1.33.

### OUTCOMES

Overall, 6 (2.2%) neonates died. Four (4.7%) out of the 85 neonates with abnormal CUS findings died, while 3 (10%) out of the 30 neonates with clinical diagnosis of meningitis died, Table 5. Among the 9 infants with abnormal CSF findings, only 1 (11.1%) died; while there was no mortality recorded among those with normal CSF findings. There was a significant difference in the outcome accross the 4 categories of neonates, (Chi=18.898, p<0.001). The neonates with meningitis, who were diagnosed both clinically and with CUS were the least likely to survive in the study, OR 0.057 (95%CI 0.01, 0.37), p=0.003; mortality rate of 16.7%. Five out of the 6 deaths were boys.

**Table 5:** Categories of Neonates and Outcomes.

|  |  | Outcome |  | Total (%) |
| --- | --- | --- | --- | --- |
|  |  | Died (%) | Discharged (%) |  |
| Categories | Pos CUS and Clinical Diagnosis (NSM) | 3 (16.7) | 15 (83.3) | 18 (100) |
|  | Pos CUS diagnosis (NSM) | 1 (1.5) | 66 (98.5) | 67 (100) |
|  | Pos Clinical diagnosis (NSM) | 0 (0) | 12 (100) | 12 (100) |
|  | Sepsis (NS) | 2 (1.1) | 174 (98.9) | 176 (100) |
| Total |  | 6 (2.2) | 267 (97.8) | 273 (100) |

## Discussion

This study examined the utility of cranial ultrasonography (CUS) for the early diagnosis, management, and outcome improvement of neonatal meningitis (NM) at the Neonatal Centre of Excellence of the University Teaching Hospital in Zambia. The findings demonstrate several important insights and implications for clinical practice in resource-limited settings.

### Summary of Study Result

Two hundred and seventy-three term mother-neonate pairs were recruited and divided into 4 diagnostic subgroups. These 4 groups were those with: positive CUS diagnosis for NSM (24.5%), positive clinical diagnosis for NSM (4.4%), positive CUS & clinical diagnosis for NSM (6.6%) and those with negative CUS & clinical diagnosis for meningitis (64.5%). Late-onset sepsis was documented in only 31.9% of neonates with Meningitis. Overall, the median random blood sugar level at admission was 5mmol/l (range 1-97mmol/l; IQR 4, 6mmol/l); duration of hospitalization was 11days (range 2-45days; IQR 6,16) and chronological age of the neonate 13days (range 1-180days; IQR 7, 21). The age of the neonates at admission (p=0.002) and their duration of hospitalization (p=0.042) were significantly different across the 4 categories of neonates in the study. Neonates with Meningitis were older in age and stayed longer on the ward than those with Sepsis; while clinical diagnosis of meningitis was made at a later age and was associated with a shorter duration of hospitalization than CUS diagnosed meningitis. The head circumference of neonates with positive CSF findings for NSM were significantly smaller by 1.79cm (95% CI -3.37, -0.21) than those with normal CSF, p=0.03. Prominent sulci, hyperechoic brain lesions, ventriculitis, and lateral ventriculomegaly were the most common abnormal findings on CUS. **Prevalence of Near-Miss cases** of NSM was at least 27.6%. A neonate with clinical diagnosis of meningitis was 3.94 times (95% CI 1.80, 8.62; p<0.001) more likely to have abnormal cranial ultrasound. **Percent Agreement** between Clinical and CUS diagnosis for NSM was 71.1%. Notably, only 12 (4.2%) of neonates underwent lumbar puncture/ventricular tap, underlining barriers to CSF-based diagnosis in this setting. Nine (75%) out of the 12 CSF reports was positive for meningitis; and the majority (55.6% [5]) of the positive CSF report belonged to the category of neonates with sepsis only. Two of these CSF samples were straw colored, 3 were cloudy, 2 were blood stained and 5 were clear. CSF culture yielded no organism in all the 12 CSF samples. One of the cloudy CSF samples yielded *Toxoplasma Gondii* by CSF serology. Ten (83.3%) out of the 12 neonates with documented CSF results were referred to UTH from other health facilities.

Sensitivity of clinical diagnosis of NM was 22.2%, Specificity was 33.3%. Positive Predictive value was 50%. Negative predictive value was 12.5%. Likelihood Ratio was 0.33. Sensitivity of CUS was 44.4%; Specificity of CUS was 66.7%. Positive Predictive value was 80%. Negative predictive value was 28.6%. Likelihood Ratio was 1.33.

Mortality rate was 2.2%. Mortality (16.7%) was highest amongst those with both clinical and CUS diagnosis of meningitis; 11.1% in those with abnormal CSF; 4.7% among those with CUS diagnosis of meningitis and none among those with clinical diagnosis of meningitis.

### Diagnostic Value of Cranial Ultrasound

A key observation from the data is that CUS identified a substantially higher number of neonates with abnormal findings suggestive of meningitis (31.1%) compared to clinical diagnosis alone (11%). This resulted in a near-miss rate of at least 27.6% (further evidence revealed by the higher numbers of positive CSF among those with sepsis only) reflecting a large proportion of mild or early cases of NSM that would have been overlooked, if relying solely on clinical presentation. The agreement between clinical and CUS diagnosis was significantly modest (71%, Kappa = 0.180), highlighting the limitations of clinical judgment in the neonatal population, where symptoms are often non-specific.

According to literature, CUS can recognize early signs of NSM with high accuracy (approximately 65%), within a week of onset of infection, whether symptomatic or not and almost 100% accuracy in identifying late signs of acute NSM presenting more than a week after onset of infection or in those who are severely symptomatic.^2,5^ Patel et al. (2019)^3^ also documented abnormal CUS findings in 81.67% (21 of 26) infants with suspected meningitis, higher than the 60% (18/30) obtained in this study.

Prominent sulci, hyperechoic brain lesions, ventriculitis, and lateral ventriculomegaly were the most common abnormal findings on CUS, consistent with previous literature.^5^ The most common and earliest finding on CUS were echogenic sulci in 92.6%, ventriculomegaly (77.8%) and parenchymal abnormalities in 62.9% of these neonates in Nakazibwe et al. (2009)’ s Uganda Mulago Hospital study.^5^ Other prognostic determining findings were ventriculitis and cerebral edema.^5^ In this Mulago study, the utility of CUS was much higher (sensitivity of 85% and specificity of 74%) than what was obtained in this current study. Perhaps this discrepancy in CUS utility, can be explained by the low CSF uptake and non-inclusion of the report on the Doppler component of CUS in this current paper. CUS is extremely useful in evaluating ventriculitis and intraventricular contents, hence, several authors have recommended that CUS be performed in every infant with clinical suspicion of sepsis and meningitis.^9, 3^

The reasonable sensitivity and specificity of CUS, seen in this study, support its use as an adjunct or alternative to lumbar puncture, especially where parental consent for LP is low or LP is technically challenging or contraindicated.

### Low uptake of Lumbar puncture and Ventricular tap

The present investigation revealed a low adoption rate of 4.4% for LP or VT, a figure consistent with findings in other research. While this study did not delve into the causes of this limited uptake, Masood et al. (2023) identified factors such as illiteracy, paternal decision-making within patriarchal societal structures, peer influence, and insufficient knowledge as reasons for caregivers declining consent for such invasive procedures, even after receiving comprehensive prior information.^22^

### Irreplaceability of CSF Analysis

All the 12 CSF reports were culture negative, probably due to prior antibiotics exposure as majority were referred from other hospitals and our laboratories were unable to perform molecular tests on the CSF. The majority of the positive CSF reports, specifically 55.6% [5], were associated with neonates diagnosed solely with sepsis. This finding underscores the indispensable role of CSF analysis in diagnosing meningitis. Nevertheless, the utilization of lumbar puncture or ventricular tap in this investigation was notably low, at a mere 4.4%. Caregivers were reluctant to provide consent for this invasive procedure, even subsequent to receiving comprehensive information. A potential strategy to address this suboptimal uptake of lumbar puncture/ventricular tap procedures may involve performing them on all neonates presenting with suspected sepsis upon arrival at the emergency room, a time when caregivers are typically highly motivated to collaborate with healthcare professionals to ensure optimal patient care.^1,2,3^

### Correlation with CSF and CUS/Clinical Diagnosis

CUS findings showed stronger (twice stronger) correlation with CSF results than clinical diagnosis. The sensitivity and specificity of CUS (44.4% and 66.7%, respectively) and its higher positive predictive value (80%) compared to clinical evaluation alone further reinforce its diagnostic utility. These CUS utility values were lower than those documented in a cross-sectional study of 227 infants, with acute bacterial infection in Mulago Hospital, Uganda [2009], where CUS was found to have a sensitivity of 85% and specificity of 74.1%, when compared with CSF culture.^5,23^ However, culture-negative CSF results, possibly related to prior antibiotic exposure or viral etiologies, and low rates of CSF analysis, in this current study limited the gold standard comparison.

### Clinical and Demographic Predictors

Rate of clinical NSM in suspected sepsis was 11% in this current study; lower than the 19.3% found in the Ethiopia study.^6,7^ No significant differences were found in most demographic or clinical variables across diagnostic categories, except for age at admission and duration of hospitalization. Those with only clinical NSM had a shorter hospital stay compared with those with CUS-NSM; this shorter stay may be due to uncertainty surrounding the diagnosis of NSM based on clinical assessment alone. Notably, subgroup of neonates with clinical & CUS diagnosis of NSM had the lowest hemoglobin and oxygen saturation, and highest pain scores, potentially reflecting greater disease severity.

### Outcome and Prognosis

The mortality rate observed in this investigation, which was 2.2% among 273 neonates admitted for sepsis, demonstrated a lower incidence compared to the 7.7% recorded for meningitis. This finding emerged from a prospective observational study conducted at a tertiary care neonatal unit in Northern India between February 2020 and July 2021,**^24^** which enrolled neonates presenting with clinical indications of sepsis. Furthermore, this rate was lower than the 6.8% mortality reported in a retrospective study performed in Turkey in 2015.**^25^**A potential explanation for this disparity may be attributed to the exclusive inclusion of term neonates in the present study.

Only 1 of the 6 neonates that died had positive CSF for meningitis in this present study, whereas 4 of those mortalities had positive CUS findings for NSM. Mortality was highest among the sub-group of neonates with clinical & CUS-diagnosed NSM (16.7%), and none among those with clinical NSM alone. The use of CUS enabled prompt identification of high-risk cases, potentially allowing for more timely intervention. The overall mortality rate (2.2%) and the observed association between abnormal CUS and poorer outcomes underscore the importance of accurate and early diagnosis. Therefore, even amongst those that survived in subgroup of Clinical & CUS diagnosed meningitis, would need close longterm follow up post-discharge searching for symptoms and signs of neurologic sequaelae.

## Limitations and Strength

The study was limited by low rates of LP and CSF analysis, potential operator dependency of CUS, its inability to detect subtle or early changes in some cases and non-repeat of CUS procedure, especially in cases where initial CUS finding was not suggestive of meningitis. Additionally, the study was conducted in a single center, which may affect generalizability. Nevertheless, it provides valuable local evidence in a context where data are scarce and decision-making is often hampered by lack of diagnostic resources.

## Conclusion

Cranial ultrasonography is a valuable tool for the prompt detection and management of NSM in settings where lumbar puncture and advanced laboratory diagnostics are limited. Its non-invasive nature, portability, and ability to identify key features of NSM make it a practical adjunct to clinical evaluation. The efficacy of CUS demonstrated a moderate impact, limitations in detecting early stages of NSM and the presence of positive CSF among neonates with sepsis only within this investigation, further reaffirms the irreplaceability of CSF analysis in the diagnosis of meningitis. However, the acquisition of caregiver consent for lumbar puncture or ventricular tap, along with the reporting of their laboratory outcomes, were exceedingly insufficient. Nevertheless, the integration of clinical assessment and CUS findings for diagnosing meningitis emerged as possessing greater clinical significance in contexts characterized by limited resources. The study demonstrated that CUS identifies a significant proportion of cases missed by clinical assessment alone, and its findings are more closely aligned with CSF abnormalities than clinical diagnosis.

Routine implementation of CUS for neonates with suspected sepsis or meningitis (while exploring measures to improve on uptake of CSF analysis) could improve prompt and early diagnosis, guide appropriate therapy, and potentially reduce mortality and long-term neurological sequelae, particularly in resource-constrained environments. Training and capacity building for sonographers and clinicians in neonatal neuroimaging are recommended to maximize the benefits of this modality. Further multi-center research and long-term follow-up studies are warranted to consolidate these findings and inform policy and clinical guidelines.

## Data Availability

All data produced in the present study are available upon reasonable request to the authors

## Authors Contributions

Dr Ogah Adenike Oluwakemi: Corresponding Author; Conceptualization (lead), writing – original draft (lead), writing-review and editing (equal), formal analysis (lead). Prof David Hamer guided on the topic, supervised the concept and proposal development and manuscript review and editing.

## Acknowledgements

The authors are extremely grateful to the participants involved in this study, the staff of A08 ward (NCOE), Management of UTH-Children Hospital, Professor Pradeep Suryiwanyanshi, Dr Fwemba, Dr Teddy Mandoka, Dr James-Aaron Ogbole Ogah and Mr Kasongo Geofrey for their immense contributions to this study.

---

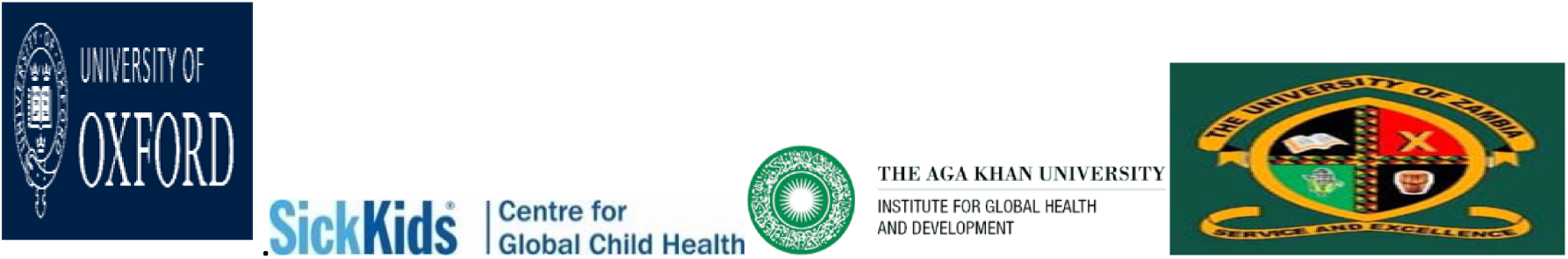

